# Considerations in the deployment of novel universal vaccines against epidemic and pandemic influenza

**DOI:** 10.1101/19002485

**Authors:** N Arinaminpathy, S Riley, W.S Barclay, C Saad-Roy, B Grenfell

## Abstract

There is increasing interest in the development of new, ‘universal’ influenza vaccines (UIV) that - unlike current vaccines - are effective against a broad range of seasonal influenza strains, as well as against novel pandemic viruses. Even where these vaccines do not block infection, they can moderate clinical severity, reducing morbidity and mortality while potentially also reducing opportunities for transmission. Previous modelling studies have illustrated the potential epidemiological benefits of UIVs, including their potential to mitigate pandemic burden. However, these new vaccines could shape population immunity in complex ways. Here, using mathematical models of influenza transmission, we illustrate two types of unintended consequences that could arise from their future deployment. First, by reducing the amount of infection-induced immunity in a population without fully replacing it, a seasonal UIV programme may permit larger pandemics than in the absence of vaccination. Second, the more successful a future UIV programme is in reducing transmission of seasonal influenza, the more vulnerable the population could become to the emergence of a vaccine-escape variant. These risks could be mitigated by optimal deployment of any future UIV vaccine: namely, the use of a combined vaccine formulation (incorporating conventional as well as multiple universal antigenic targets), and by achieving sufficient population coverage to compensate for reductions in infection-induced immunity. As early candidates of UIVs approach advanced clinical trials, there is a need to monitor their characteristics in such a way that is focused on their potential impact. This work offers a first step in this direction.

## Introduction

Vaccination has been a mainstay of influenza control since the 1940s ^1^. It has had considerable impact in averting mortality, particularly in risk groups such as the elderly, and may also have contributed towards reducing opportunities for transmission ^2–4^. Current ‘strain-matched’ vaccines raise immunity to the viral surface protein haemagglutinin (HA), particularly its immunodominant ‘head’ region ^5^. This immunity can be effective as long as it is closely matched with circulating influenza strains. Inactivated influenza vaccines typically have a ‘vaccine efficacy’ (measured using the relative risk of influenza illness amongst vaccinated vs non-vaccinated individuals) of around 60% ^6^. Recent years have also seen the deployment of live attenuated influenza vaccines ^7^, which – along with the immunodominant HA – may also raise immunity to additional viral antigens, by permitting limited rounds of replication within immunised hosts. However, there have been recent drops in live vaccine efficacy in the USA ^8^. Such changes may have arisen from poor HA matching between circulating and immunising strains, from a failure of the attenuated virus to replicate at the site of immunisation, or from a build-up of vaccine-derived immunity in target age groups ^9^.

The HA head is the most variable viral component, showing ongoing change in the human population over time as a result of population immunity ^10^. As a result current influenza vaccines have to be updated regularly, to remain effective in the face of viral evolution ^11^. Moreover, they are of limited use in the event of a pandemic (the emergence of a transmissible, antigenically novel, influenza virus in the human population). Pandemics are notoriously unpredictable in their timing, as well as in the HA subtype involved. Because of their long development time, current, strain-matched vaccines are of limited use in mitigating against these risks ^12^.

In light of these challenges, there has been increasing interest in new vaccines that can focus ‘cross-protective’ immunity against alternative viral components, that may be more conserved across HA strains and subtypes, and more constrained than HA, in their capacity for antigenic evolution ^11,12^. Some potential vaccine targets are listed in table 1. Candidates include the ‘stem’ region of the HA protein ^13^, which is more conserved than the variable ‘head’ region, as well as proteins inducing T-cell immunity ^14^, including matrix structural proteins (M1, M2) and viral nucleoprotein (NP). Recombinant and viral vector technology allows the formulation of vaccines that focus immunity on conserved antigenic targets, bypassing the immunodominant HA head region altogether. Several such ‘universal’ influenza vaccines (UIVs) are currently in development ^15^; they may offer qualitatively new opportunities for influenza control, raising the prospect of routine vaccine programmes that do not need to be updated as often, while also promoting immunity in the population that would protect against a novel pandemic virus.

**Table 1.** Summary of different immune targets for influenza vaccines. Amongst current influenza vaccines, ‘inactivated’ vaccines focus on HA1 immunity (top row), while ‘live attenuated’ vaccines allow a round of viral replication, and may therefore raise both HA-specific and T-cell immunity. However, their heterosubtypic protection is unclear. The lower two rows correspond to strategies being pursued for the development of new, ‘universal’ vaccines (we adopt the scenario in the bottom row for the purpose of the current work).

| Antigenic target | Type of immunity | Breadth of immunity | Relevant sources |
| --- | --- | --- | --- |
| Haemagglutinin (HA1, head region) | Sterilising | Strain-specific, and immunodominant; seasonal vaccines need to be updated regularly, and are not effective against novel pandemic strains | 6,37 |
| Haemagglutinin (HA1, conserved epitopes in head region) | Sterilising | Vaccine targets identified through computational methods, and may offer broad, within-subtype protection | 38,39 |
| Haemagglutinin (HA2, stalk region) | Sterilising | Broad protection within and across subtypes (animal models): could offer pandemic protection in humans | 40–42 |
| T-cell antigens: e.g. matrix proteins (M1 & M2), nucleoprotein (NP) | Non-sterilising, but could reduce clinical severity, and potentially infectiousness, by limiting viral load | Broad protection within and across subtypes (animal models): could offer pandemic protection in humans | 43–45 |

Different vaccine formulations also offer different types of immune protection, for example with HA-based vaccines providing ‘sterilising’ protection, that can halt infection in its early stages ^16^. By contrast, vaccines based on T-cell antigens require some viral replication to occur, in order for these antigens to be processed and made available for immune recognition ^17^. Such vaccines thus do not offer sterilising immunity, but instead can moderate the clinical course of infection, potentially also reducing onward transmission ^18^. Table 1 summarises some examples of antigenic targets against influenza, and their potential use in vaccine formulations.

Previous evidence from disease-dynamic models illustrated the possible population benefits of mass deployment of UIVs, for example that such vaccines could mitigate the impact of novel pandemic viruses. Moreover, over several years, modelling illustrated how sustained vaccination with UIVs could slow the HA evolution of seasonal influenza, by simultaneously suppressing the generation of, and selection pressure for, new antigenic variants ^19^. Nonetheless, these studies had two main limitations: first, they were not specific to a particular epidemiological setting, aiming instead to illustrate underlying dynamics in a parsimonious but generalisable way. Second, they did not aim to capture immune complexities in the population, nor how these might be shaped by a UIV.

However, population immunity to influenza is complex and incompletely understood, raising the possibility of unintended consequences from mass vaccination with UIVs. For example, despite the antigenic novelty associated with pandemic viruses, recent work showed that pre-existing T-cells to CD8 epitopes could nonetheless moderate the clinical severity of disease ^7^. In the present work we aim to examine the epidemiological implications of these dynamics. We address the questions: could a UIV programme affect influenza transmission in adverse ways? How might these risks be mitigated, and what are the critical vaccine characteristics that would need to be monitored to do so?

We use simple mathematical models of influenza immunity and transmission, focusing on the USA, which currently has the world’s largest coverage of seasonal influenza vaccination ^20^. A key advantage of this setting is that we are able to build on previous work ^21^, that captured vaccine efficacy and epidemiological impact of existing influenza vaccination efforts in the USA, over the last decade: we adopt these calibrated models to project the potential impact of UIVs in this setting. We note that the aim of the present study is to identify potential scenarios rather than to make accurate quantitative predictions. We use simple models to draw attention to: (i) examples of immunological and epidemiological dynamics that could result in adverse effects of UIV programmes, and (ii) how these effects might be overcome. In setting out these hypothetical scenarios, this work is also a first step in identifying the key uncertainties that need to be addressed in the course of vaccine development, in order to mitigate the potential for unintended consequences.

## Results

We build on a model previously developed to capture the impact of seasonal influenza vaccination in the USA ^21^ (see Methods and materials). The model incorporates two types of immunity: (i) HA-specific immunity, which reduces susceptibility to infection. Acquired either through past exposure to infection or through effective strain-matched vaccination. (ii) Cross-protective immunity, assumed to be independent of HA-specific immunity, and likewise acquired either through past infection or through a UIV. We assume that this immunity is heterosubtypic: that is, offering protection across different subtypes. Moreover, we assume that cross-protective immunity does not affect susceptibility to infection, but rather limits viral load during the course of infection, thus reducing infectiousness. This would be consistent, for example, with a UIV targeting T-cell antigens ^22^; however, we note that HA-stem antibodies would be expected to offer some protection against infection ^23,24^ (Table 1).

Wide deployment of an effective UIV could allow sustained control of epidemic influenza without need for being updated as frequently as current vaccines. Figure 1 shows the series of influenza seasons in the USA initially modelled in ref. ^21^, illustrating the epidemic sizes that would have occurred if each dose of strain-matched vaccine had been replaced by a dose of UIV (i.e. with identical vaccination coverage). Dots show the minimum levels of vaccine efficacy that are needed, for a UIV to achieve smaller epidemic sizes than those that resulted from the strain-matched vaccination programme (for UIVs, we denote ‘efficacy’ as the percentage drop in transmission potential arising from vaccination). These minimum levels of UIV efficacy are consistent with levels of VE that are reported in practice, for strain-matched vaccines. Importantly for influenza, the basic reproduction number (R0) is typically only 1 – 2 ^25^. It is therefore possible, even for a vaccine with only modest effectiveness, to have a substantial impact on influenza transmission at sufficiently high coverage ^19^.

**Figure 1.**
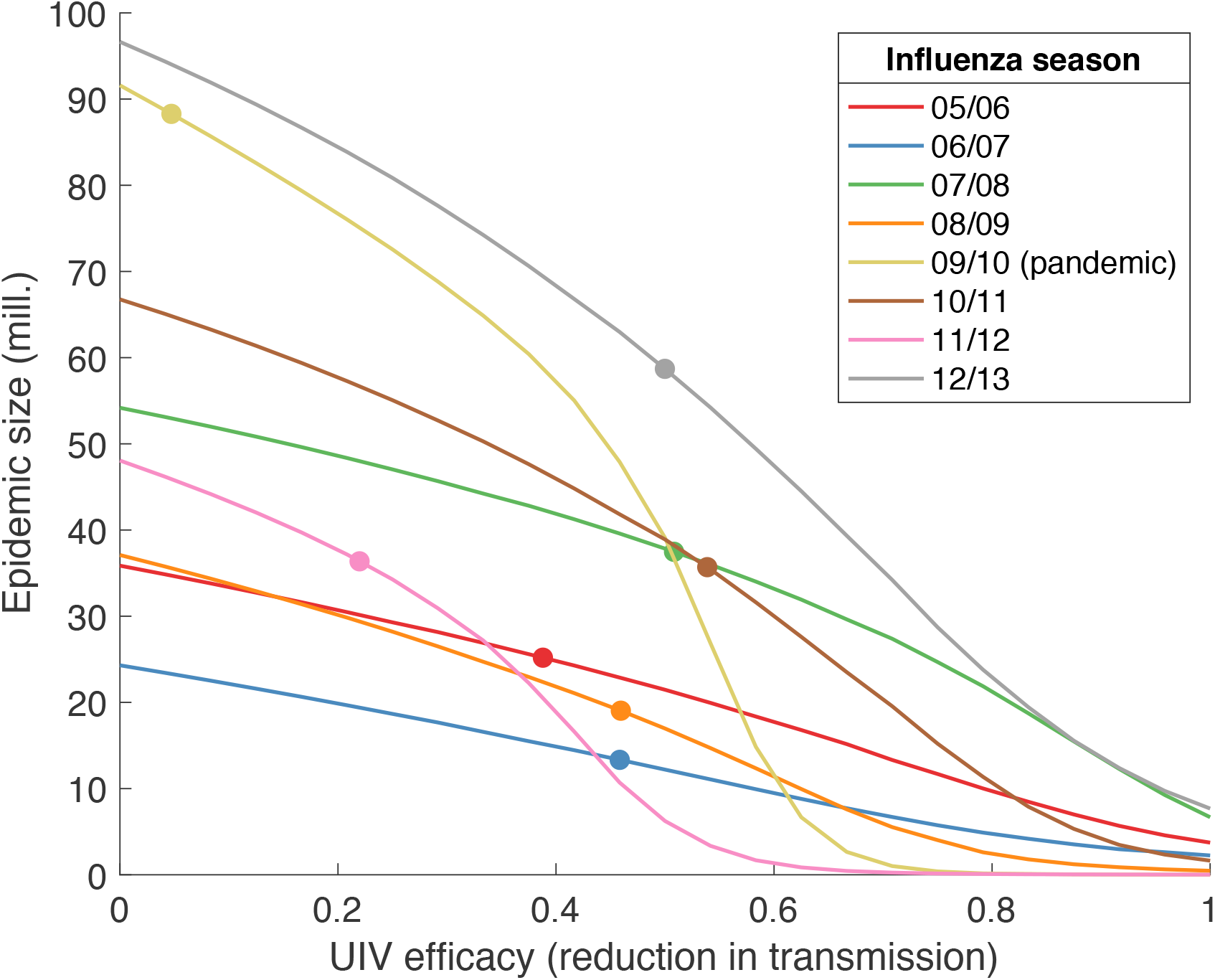
The potential impact of universal vaccination on past seasonal epidemics in the US. Using a model calibrated to historical seasonal epidemics in the US, the figure shows the epidemic sizes that would have occurred if each dose of the conventional vaccine were replaced by a UIV (universal influenza vaccine), one that offers no reduction in susceptibility, but that lowers transmission potential by a factor *c* (x-axis). Dots on lines mark the actual epidemic sizes that occurred, under conventional vaccination, for each season: a UIV with efficacy greater than indicated by these points would therefore outperform the conventional vaccines used at the time.

Moreover, notably in the case of the 2009 H1N1 pandemic, it was not possible to deploy appropriate conventional vaccines sufficiently early to mitigate the pandemic. In this case the timely deployment of even a modestly effective cross-protective vaccine could avert millions of cases.

Having illustrated these potential benefits of a UIV, we now explore their potential adverse effects. We focus on the implications of routine seasonal vaccination with a UIV. In particular, we perform the analysis illustrated schematically in Figure 2: we simulate a population that is subject to seasonal UIV vaccination, and subsequently exposed to a 2012/13-like season. Tracking population immunity following these combined immunisation events (the routine UIV programme and the seasonal epidemic), we then assess the vulnerability of the population to different types of subsequent immune escape.

**Figure 2.**
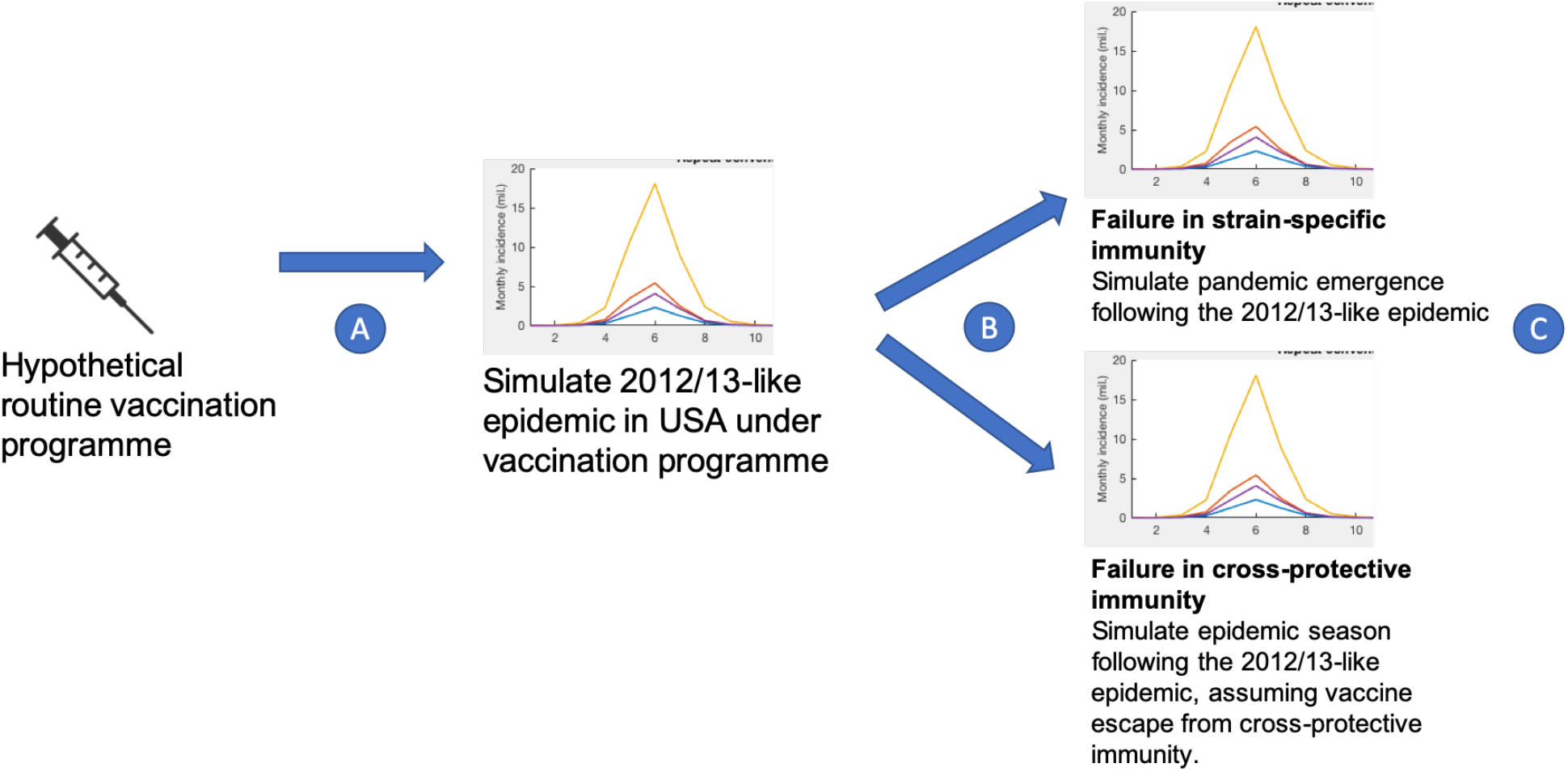
Schematic of the overall modelling approach. We focus on the use of UIVs in routine seasonal influenza vaccination. At stage (B) in the figure, population immunity depends on both the vaccination programme at stage (A), and the ensuing seasonal epidemic. However, the latter can be heavily influenced by the former; we use a dynamical transmission model to capture these relationships. We then explore the implications of immunity at stage (B) under two scenarios, illustrated on the right-hand side of the figure: (i) where a pandemic virus emerges soon after the epidemic season (upper right-hand plot), and (ii) alternatively, where a second epidemic season is caused by a variant capable of escape from cross-protective immunity (lower right-hand plot). In each scenario, we assess how final epidemic sizes at stage (C) are shaped by the choice of vaccination programme at stage (A).

We begin by examining **pandemic emergence**. First, we focus on how the routine seasonal UIV programme illustrated in Figure 2 may affect pandemic emergence: we assume there is no supplementary UIV programme mounted in direct response to the pandemic. Figure 3 shows one illustrative scenario, where a pandemic virus emerges soon after the 2012/13- like season. The figure suggests that, at low levels of coverage, a seasonal UIV programme may in fact increase the pandemic size. Figure 3B illustrates why: by bringing down seasonal incidence, the UIV programme reduces opportunities for individuals to acquire infection-induced immunity. As a result, there is an expanded pool of individuals who have neither been vaccinated nor infected, and who are thus vulnerable to the emergence of a pandemic virus. As discussed below, this result is analogous to previous work ^26^, that showed how vaccination in individuals could reduce opportunities for those individuals to acquire broadly protective immunity; Figure 3A illustrates how such a phenomenon could arise at the population level. However, the figure also illustrates that it is possible to overcome this population effect if a seasonal UIV programme has sufficient coverage. In particular, a UIV having 80% efficacy and at 75% coverage would not only interrupt transmission for the 2012/13-like season, but would also result in a mitigated pandemic, relative to the absence of vaccination.

**Figure 3.**
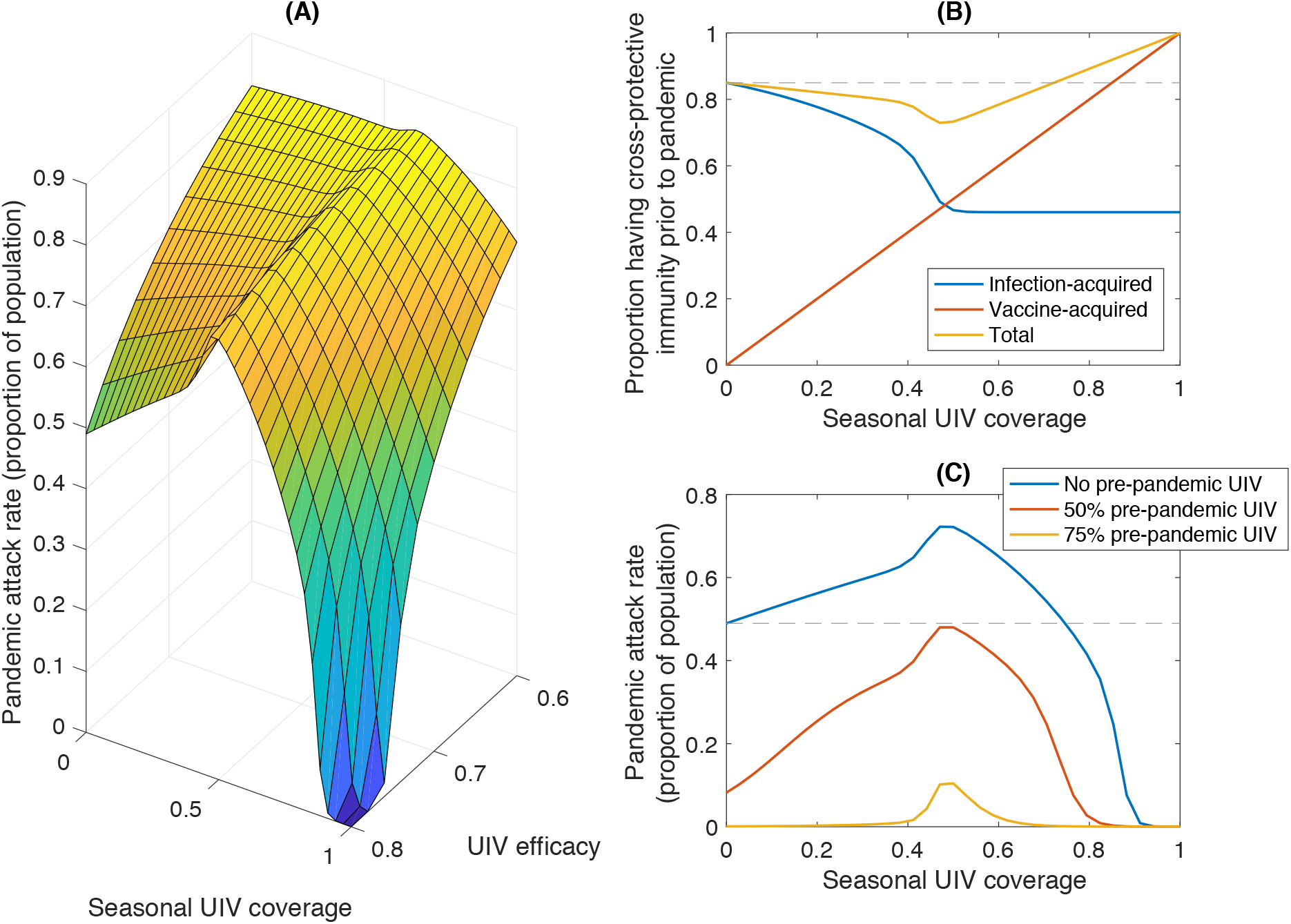
Implications of routine seasonal UIV for a population’s vulnerability to pandemic influenza. As illustrated in figure 2 we model a 2012/13-like season in the USA, simulating a scenario where conventional vaccines prior to this season were replaced by a UIV vaccine. We then simulate the emergence of a novel pandemic strain, first assuming no vaccination programme against this pandemic. Here and throughout, we define ‘efficacy’ of a UIV as the percentage drop in transmission potential arising from vaccination. (A) Pandemic size under a range of values for seasonal UIV coverage, and for UIV efficacy. Low seasonal UIV coverage, especially for an efficacious vaccine, can yield a greater pandemic than in the absence of vaccination. (B) An explanation of the behaviour in panel A, by showing separately the total amount of vaccine-derived and infection-derived immunity in the population, at a cross-section where UIV efficacy is assumed to be 80% (i.e. the ‘leading edge’ of panel A). By bringing down seasonal epidemic sizes, seasonal universal vaccination also brings down the total amount of infection-acquired immunity in the population (blue curve). At low coverage, the vaccination programme fails to compensate for this loss of immunity (yellow curve, initial decline). The dashed grey line indicates the level of cross-protective immunity in the absence of the UIV programme. The UIV programme succeeds in increasing population immunity beyond this point at a threshold coverage of approximately 75%. (C) Relaxing the assumption of no pre-pandemic vaccination, again taking the cross-section corresponding to a UIV efficacy of 80%. As in panel B, the horizontal grey line indicates the pandemic size in the absence of vaccination.

In Figure 3C we relax the assumption of no pre-pandemic vaccination. Taking as an example the case of UIV efficacy of 80% (that is, corresponding to the ‘leading edge’ of the surface in Figure 3A), Figure 3C illustrates pandemic outcomes under different scenarios for pre-pandemic UIV coverage. Overall, if seasonal UIV programmes can increase a population’s vulnerability to new pandemic viruses, panels B, C illustrate two ways in which it is possible to counteract this effect: either with sufficiently high vaccination coverage in seasonal UIV programmes, as noted above (point B), or in pre-pandemic UIV programmes (panel C).

We next examine the emergence of a seasonal virus that is a **UIV escape variant**: that is, a virus showing vaccine escape to all UIV antigenic targets, but not against strain-defining epitopes in the HA head. Owing to the plasticity of HA, such a scenario may be applicable to anti-HA1 and anti-HA2 antibodies (see Table 1). Overall, although conserved antigens typically face functional constraints that are thought to limit their diversity ^27^, the new selection pressures raised by UIVs could, in principle, promote the emergence of immune escape amongst UIV targets. Figure 4 shows the scenario where a 2012/13-like epidemic is followed by a season caused by a UIV escape variant. The figure shows two scenarios: where routine vaccination is conducted using strain-matched vaccines (panel A), and where it is instead conducted using a UIV (panel B). Under the first scenario, the two epidemics are of comparable size. Under the second scenario, although the UIV succeeds in controlling the initial epidemic, it does so at the expense of strain-specific immunity. As a result the subsequent season, associated with a UIV-escape mutant, is considerably larger than it otherwise would have been (the latter indicated by the dashed, horizontal lines for comparison). As discussed below, such risks could be mitigated by a combined vaccination strategy.

**Figure 4.**
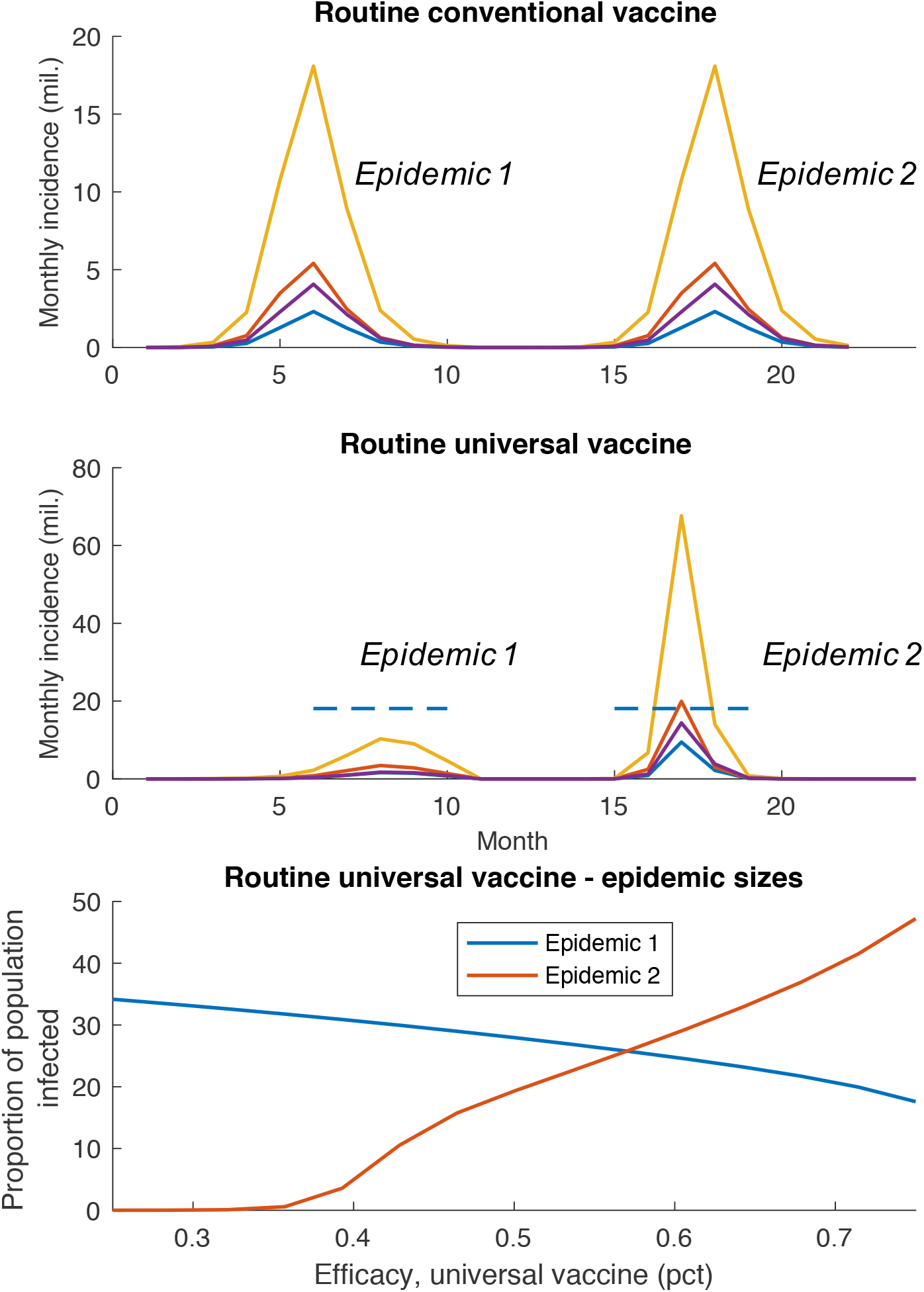
Potential impact of vaccine failure. Figures show dynamics arising from a 2012/13-like epidemic (epidemic 1), followed by a seasonal virus capable of escape from cross-protective immunity (epidemic 2). (A) With routine seasonal vaccination using a conventional (strain-matched) vaccine, epidemic sizes are unchanged by this type of immune escape. (B) With routine seasonal vaccination using a UIV, successful control of epidemic 1 can have the unintended effect of permitting a larger epidemic 2. Dashed lines show the epidemic peaks reached under a conventional vaccine (panel A), for comparison. (C) How epidemic sizes in panel B change with UIV efficacy in epidemic 1 (we assume throughout that this efficacy declines to 25% for epidemic 2, as a result of vaccine escape). The figure illustrates how improving control of seasonal influenza (blue curve) could in fact leave a population more vulnerable to an immune escape variant (red curve).

## Discussion

Population immunity makes for a complex and dynamic immune landscape. These intricacies are only increased by the different types of immunity in effect: on the one hand, transmission-blocking immunity that is focused against specific antibody targets, and on the other, more broadly acting immunity that may only be partially transmission-blocking, allowing complex immune dynamics. By focusing on specific types of immunity over others, a vaccination programme may shift the landscape of population immunity in unexpected ways, particularly in settings such as the USA, where routine coverage already exceeds 40% of the population ^20^. With the prospect of new vaccination programmes that could induce strong indirect protection (i.e. through reducing opportunities for transmission), it is important to anticipate potential adverse effects, and how to overcome them. Here we have illustrated two potential risks, corresponding respectively to the effects of compromising cross-protective immunity (Figure 3) and of compromising strain-specific immunity (Figure 4).

First, owing to the nonlinear dynamics of influenza transmission, at low coverages in routine seasonal vaccination, a UIV could in principle control seasonal epidemics, without fully compensating (at the population level) for the reduced opportunities for individuals to gain infection-induced immunity (Figure 3). Doing so would leave the population more vulnerable to a pandemic. Indeed, a similar effect has been proposed in the context of conventional vaccines: namely, that such vaccines could – by neutralising, strain-matched immunity – reduce opportunities for individuals to acquire cross-protective immunity through infection ^26^. Such effects also arise through the direct protection afforded by strain-matched vaccines; here we demonstrate ways in they could even arise indirectly (i.e. through reducing transmission) at the population level. They could, therefore, adversely affect those who have not received the vaccine.

Second, by controlling seasonal epidemics, seasonal UIV programmes may also compromise strain-specific immunity in the population: this becomes a concern in the context of viable, transmissible escape mutants to cross-protective immunity. Despite strong arguments for why certain immune targets may face functional constraints preventing them from expressing significant variation ^27,28^, the possibility of escape cannot be fully discounted ^29^. Nonetheless, a vaccine formulation combining several different antigens may well greatly reduce the risk of vaccine escape.

We also note that these risks could be mitigated to some extent, by adequate planning of a future vaccination programme: for example, Figure 3A suggests that a routine UIV programme could indeed protect against pandemics (i.e. reduce the pandemic size) if it has a sufficiently high coverage, to overcome the loss of infection-induced immunity. Figure 3A suggests a minimum coverage of 75%. This threshold is substantially greater than the standard ‘critical vaccination threshold’ required to interrupt transmission of seasonal influenza (points A and B in Figure 3) although, in practice, will depend on several factors including the relative strength and duration of immunity imparted by vaccination versus infection. Likewise in the case of compromising strain-specific immunity, the risks arising from a seasonal UIV programme would be limited by an approach that deploys conventional, strain-matched vaccines in parallel with UIVs. Overall, such an approach – for example, through a combined formulation – could mitigate strongly against the risk of vaccine escape, to either type of vaccine.

As with any modelling study, our framework has some limitations to note: first, we adopted a highly simplified representation of influenza immunity. We ignored any potential interactions between strain-specific and cross-protective immunity, for example the fact that B-cells can promote the cellular immune response, and vice versa ^30,31^. With improved data on the implications of these mechanisms for transmission, future work should explore these interactions more closely, to examine their implications for vaccine development. Additionally, for simplicity we assumed that cross-protective immunity is short-lived, and does not persist between seasons. To some extent this was necessitated by the data available, which does not allow us to estimate separately the roles of these two different arms of immunity. However, the duration of cross-protective immunity elicited by immunisation will be an important characteristic of future UIVs ^32^. There is a need for systematic studies to quantify this duration by monitoring cross-protective immunity over several years. For simplicity we assumed vaccine- and infection-induced immunity to be identical. An important question for future work is the implication of vaccine-induced immunity that is inferior to infection-induced immunity: how should future vaccination programmes compensate for any such shortfalls? What aspects of immune protection (strength, breadth, duration, etc) are most important to address? To inform such analysis, cohort studies of individuals with different exposure and vaccination history would be invaluable.

We also ignored long-term immune dynamics that could shape protection decades after exposure: two important examples are antigenic ‘imprinting’, where childhood exposure offers heterosubtypic protection against zoonotic viruses later in life ^33^, and ‘antigenic seniority’, where HA immune responses during a host’s lifetime tend to be biased towards those strains encountered early in life ^34^. Other immune complexities that we have ignored include short-lived, ‘strain-transcending’ immunity that has been proposed to explain influenza evolutionary patterns ^35^; the complex relationship between immunodominance and transmission-blocking ^36^; and the potential impact of UIVs on seasonal influenza evolution ^19^. Nonetheless, as noted above, the aim of the current work is not to provide definitive projections, but rather to illustrate the potential for population immune dynamics to give rise to unintended consequences. Addressing the limitations described above, through improved understanding of influenza immunity, would enable more robust estimates of important parameters emerging from our analysis: for example, the minimum vaccination coverage in order for a population to be protected against a pandemic (Fig.4).

In conclusion, although vaccination has offered great benefits in the control of influenza for several decades, new technologies may offer qualitatively new opportunities for influenza control; whether allowing routine seasonal vaccination programmes to adopt a stable vaccine formulation over several years, or enabling programmes that can mitigate pandemic risk on a population level. Previously we used simple models to illustrate these potential benefits (15, 28). In the current work we build on these studies, incorporating additional detail on immune dynamics to reveal some potential adverse effects of UIV programmes. However, our results also show how these effects could be effectively mitigated, through appropriate vaccine deployment. In view of the complex effects that UIVs could have on population immunity, the design of their ‘target product profiles’ would benefit significantly from population dynamics and evolutionary considerations.

## Materials and Methods

We built on a model previously developed to capture the impact of seasonal influenza vaccination in the USA. The model framework is described in detail in ref ^21^. In brief, the model is a deterministic, compartmental, age-structured framework, capturing the influenza epidemic in the USA at the national level, and allowing for different levels of prior immunity in each of the age groups.

### Modelling immunity

As described in the main text, the model incorporates two types of immunity: (i) HA-specific immunity, which reduces susceptibility to infection, and (ii) Independently acting cross-protective immunity, assumed not to affect susceptibility to infection, but rather to reduce infectiousness (Table 1). For illustration we assumed vaccine-derived immunity to elicit identical protection as infection-induced immunity (in strength and duration), but focusing solely on specific antigens, whether HA-specific (as for conventional, strain-matched vaccines) or broadly protective (as for universal vaccines). For simplicity, we focus on inactivated vaccines and not live attenuated vaccines, the former of which account for the majority of influenza vaccination in the USA. Moreover we did not aim to model the dynamics of immunity across different influenza seasons, instead treating each epidemic independently. In the absence of serology or other immunological data, it is not possible to estimate separately the roles of HA-specific or cross-protective immunity, for a given epidemic; instead we took the extreme (but simplifying) assumption that any cross-protective immunity is short-lived, and does not persist between epidemic seasons. Thus, all immunity at the beginning of the season is HA-specific: we estimated the age-specific proportions having this immunity using the data described above.

For simplicity we ignored dynamics such as ‘antigenic imprinting’, or the protection arising from childhood infection, against exposure later in life to heterosubtypic, zoonotic influenza viruses ^33^. We also ignored the potential impact of universal vaccination on influenza evolution; this is consistent with genomic analysis suggesting that the principal source of antigenic novelty for influenza is not the USA, but in South-East Asia and in the tropics ^46,47^.

### Governing equations and calibration

For a single season, we neglected births and deaths in the host population. In the following equations, we denote age groups with subscript *i*, where *i* = [1, 2, 3, 4] denote respectively 6 mo – 4 yrs, 5 – 19 yrs, 20 – 64 yrs, and >65 yrs. We denote vaccination status with the conventional (HA-based) vaccine as *j*, where *j* = [0, 1] denotes respectively unvaccinated and vaccinated individuals. Similarly we denote as *k* the vaccination status with the cross-protective vaccine. We write *S*_*ijk*_, *I*_*ijk*_, *R*_*ijk*_for the proportions of the population that are, respectively, susceptible, infected and recovered, each stratified by the age and vaccination categories *i,j, k*.

The strain-matched vaccine reduces susceptibility to infection by a proportion *p*_*i*_in age-group *i*, and cross-protective vaccination reduces infectiousness by a proportion *q*_*j*_in age-group *j*.

Governing equations for the model are:

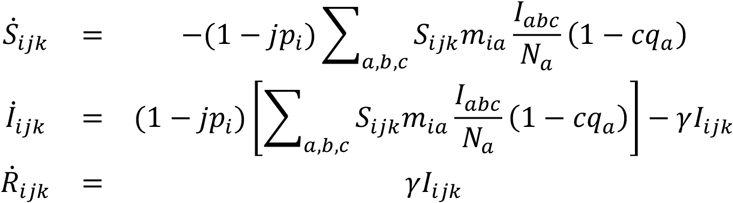

where *γ* is the per-capita rate of recovery, and (for convenience of notation) the vaccine status indices *j, c* are being used as indicator functions (i.e. their values 0, 1 being treated not only as categorical, but also as multiplying terms in these equations).

Free model parameters, to be calibrated, included: the initial susceptibility in each age group (as a result of past, strain-specific exposure); the overall rate of transmission per day; and the age-specific effect of vaccination in reducing susceptibility. Treating each influenza season independently, these parameters were estimated using the following data sources: virologically confirmed influenza hospitalisations in each month; age-specific estimates of vaccine efficacy (VE) for each season; and estimates of monthly, age-specific vaccine coverage in each season. Additionally, the model is informed by ‘multipliers’ estimated by the US Centers for Disease Control and Prevention (CDC), which relate hospitalisations to the incidence of symptomatic illness. In ref ^21^ we describe in detail how the model was fitted to this data using Bayesian methods. In the present work, uncertainty estimates are less critical than in ref. ^21^. For the simulations we therefore selected the best fitting parameter set for the 2012/13 season, i.e. that maximising the posterior density constructed as in ref 21.

## Data Availability

This manuscript does not generate new primary data, and instead uses mathematical modelling to analyse data already available in the literature

## References

1. Treanor, J. Influenza Vaccine — Outmaneuvering Antigenic Shift and Drift. N. Engl. J. Med. 350, 218–220 (2004).

2. Piedra, P. A. et al. Herd immunity in adults against influenza-related illnesses with use of the trivalent-live attenuated influenza vaccine (CAIV-T) in children. Vaccine 23, 1540–1548 (2005).

3. Rudenko, L. G. et al. Efficacy of live attenuated and inactivated influenza vaccines in schoolchildren and their unvaccinated contacts in Novgorod, Russia. J. Infect. Dis. 168, 881–7 (1993).

4. Monto, A. S., Davenport, F. M., Napier, J. A. & Francis, T. Modification of an outbreak of influenza in Tecumseh, Michigan by vaccination of schoolchildren. J. Infect. Dis. 122, 16–25

5. Fitch, W. M., Bush, R. M., Bender, C. A. & Cox, N. J. Long term trends in the evolution of H(3) HA1 human influenza type A. Proc. Natl. Acad. Sci. U. S. A. 94, 7712–8 (1997).

6. Osterholm, M. T., Kelley, N. S., Sommer, A. & Belongia, E. A. Efficacy and effectiveness of influenza vaccines: a systematic review and meta-analysis. Lancet Infect. Dis. 12, 36–44 (2012).

7. Sridhar, S., Brokstad, K. A. & Cox, R. J. Influenza Vaccination Strategies: Comparing Inactivated and Live Attenuated Influenza Vaccines. Vaccines 3, 373–89 (2015).

8. Flannery, B. et al. Interim estimates of 2013-14 seasonal influenza vaccine effectiveness - United States, February 2014. MMWR. Morb. Mortal. Wkly. Rep. 63, 137–42 (2014).

9. Singanayagam, A., Zambon, M., Lalvani, A. & Barclay, W. Urgent challenges in implementing live attenuated influenza vaccine. Lancet Infect. Dis. 18, e25–e32 (2018).

10. Smith, D. J. et al. Mapping the Antigenic and Genetic Evolution of Influenza Virus. Science (80-.). 305, 371–376 (2004).

11. Carrat, F. & Flahault, A. Influenza vaccine: The challenge of antigenic drift. Vaccine 25, 6852–6862 (2007).

12. Fedson, D. S. Pandemic Influenza and the Global Vaccine Supply. Clin. Infect. Dis. 36, 1552–1561 (2003).

13. Wei, C.-J. et al. Induction of Broadly Neutralizing H1N1 Influenza Antibodies by Vaccination. Science (80-.). 329, 1060–1064 (2010).

14. Soema, P. C., van Riet, E., Kersten, G. & Amorij, J.-P. Development of cross-protective influenza a vaccines based on cellular responses. Front. Immunol. 6, 237 (2015).

15. Erbelding, E. J. et al. A Universal Influenza Vaccine: The Strategic Plan for the National Institute of Allergy and Infectious Diseases. J. Infect. Dis. 218, 347–354 (2018).

16. Waffarn, E. E. & Baumgarth, N. Protective B Cell Responses to Flu—No Fluke! J. Immunol. (2011). doi:10.4049/jimmunol.1002090

17. Thomas, P. G., Keating, R., Hulse-Post, D. J. & Doherty, P. C. Cell-mediated protection in influenza infection. Emerg. Infect. Dis. 12, 48–54 (2006).

18. Sridhar, S. Heterosubtypic T-Cell Immunity to Influenza in Humans: Challenges for Universal T-Cell Influenza Vaccines. Front. Immunol. (2016). doi:10.3389/fimmu.2016.00195

19. Arinaminpathy, N. et al. Impact of cross-protective vaccines on epidemiological and evolutionary dynamics of influenza. Proc. Natl. Acad. Sci. U. S. A. 109, (2012).

20. Centers for Disease Control and Prevention (CDC). nfluenza vaccine coverage estimates, 2017 - 2018 season. Available at: https://www.cdc.gov/flu/fluvaxview/1718season.htm. (Accessed: 18th November 2018)

21. Arinaminpathy, N. et al. Estimating Direct and Indirect Protective Effect of Influenza Vaccination in the United States. Am. J. Epidemiol. 186, (2017).

22. Epstein, S. L. & Price, G. E. Cross-protective immunity to influenza A viruses. Expert Rev. Vaccines 9, 1325–1341 (2010).

23. Ekiert, D. C. et al. Antibody Recognition of a Highly Conserved Influenza Virus Epitope. Science (80-.). 324, 246–251 (2009).

24. Dreyfus, C., Ekiert, D. C. & Wilson, I. A. Structure of a classical broadly neutralizing stem antibody in complex with a pandemic H2 influenza virus hemagglutinin. J. Virol. 87, 7149–54 (2013).

25. Biggerstaff, M., Cauchemez, S., Reed, C., Gambhir, M. & Finelli, L. Estimates of the reproduction number for seasonal, pandemic, and zoonotic influenza: a systematic review of the literature. BMC Infect. Dis. 14, 480 (2014).

26. Bodewes, R. et al. Annual vaccination against influenza virus hampers development of virus-specific CD8+ T cell immunity in children. J. Virol. 85, 11995–2000 (2011).

27. Wu, N. C. & Wilson, I. A. A Perspective on the Structural and Functional Constraints for Immune Evasion: Insights from Influenza Virus. J. Mol. Biol. 429, 2694–2709 (2017).

28. Berkhoff, E. G. M. et al. Functional Constraints of Influenza A Virus Epitopes Limit Escape from Cytotoxic T Lymphocytes. J. Virol. 79, 11239–11246 (2005).

29. Doud, M. B., Lee, J. M. & Bloom, J. D. How single mutations affect viral escape from broad and narrow antibodies to H1 influenza hemagglutinin. Nat. Commun. 9, 1386 (2018).

30. Alam, S., Knowlden, Z. A. G., Sangster, M. Y. & Sant, A. J. CD4 T cell help is limiting and selective during the primary B cell response to influenza virus infection. J. Virol. 88, 314–24 (2014).

31. León, B., Bradley, J. E., Lund, F. E., Randall, T. D. & Ballesteros-Tato, A. FoxP3+ regulatory T cells promote influenza-specific Tfh responses by controlling IL-2 availability. Nat. Commun. 5, 3495 (2014).

32. Subramanian, R., Graham, A. L., Grenfell, B. T. & Arinaminpathy, N. Universal or Specific? A Modeling-Based Comparison of Broad-Spectrum Influenza Vaccines against Conventional, Strain-Matched Vaccines. PLoS Comput. Biol. 12, (2016).

33. Gostic, K. M., Ambrose, M., Worobey, M. & Lloyd-Smith, J. O. Potent protection against H5N1 and H7N9 influenza via childhood hemagglutinin imprinting. Science 354, 722–726 (2016).

34. Lessler, J. et al. Evidence for Antigenic Seniority in Influenza A (H3N2) Antibody Responses in Southern China. PLoS Pathog. 8, e1002802 (2012).

35. Ferguson, N. M., Galvani, A. P. & Bush, R. M. Ecological and immunological determinants of influenza evolution. Nature 422, 428–433 (2003).

36. Angeletti, D. & Yewdell, J. W. Is It Possible to Develop a “Universal” Influenza Virus Vaccine? Cold Spring Harb. Perspect. Biol. 10, a028852 (2018).

37. Belongia, E. A. et al. Variable influenza vaccine effectiveness by subtype: a systematic review and meta-analysis of test-negative design studies. Lancet Infect. Dis. 16, 942–951 (2016).

38. Thompson, C. P. et al. A naturally protective epitope of limited variability as an influenza vaccine target. Nat. Commun. (2018). doi:10.1038/s41467-018-06228-8

39. Carter, D. M. et al. Design and Characterization of a Computationally Optimized Broadly Reactive Hemagglutinin Vaccine for H1N1 Influenza Viruses. J. Virol. (2016). doi:10.1128/jvi.03152-15

40. Impagliazzo, A. et al. A stable trimeric influenza hemagglutinin stem as a broadly protective immunogen. Science (80-.). 349, 1301–1306 (2015).

41. Margine, I. et al. Hemagglutinin stalk-based universal vaccine constructs protect against group 2 influenza A viruses. J. Virol. 87, 10435–46 (2013).

42. Krammer, F. & Palese, P. Influenza virus hemagglutinin stalk-based antibodies and vaccines. Curr. Opin. Virol. 3, 521–530 (2013).

43. Wang, W. et al. Protective Efficacy of the Conserved NP, PB1, and M1 Proteins as Immunogens in DNA- and Vaccinia Virus-Based Universal Influenza A Virus Vaccines in Mice. Clin. Vaccine Immunol. 22, 618–630 (2015).

44. Price, G. E., Lo, C.-Y., Misplon, J. A. & Epstein, S. L. Mucosal immunization with a candidate universal influenza vaccine reduces virus transmission in a mouse model. J. Virol. 88, 6019–30 (2014).

45. Gilbert, S. C. T-cell-inducing vaccines - what’s the future. Immunology 135, 19–26 (2012).

46. Russell, C. A. et al. The Global Circulation of Seasonal Influenza A (H3N2) Viruses. Science (80-.). 320, 340–346 (2008).

47. Lemey, P. et al. Unifying viral genetics and human transportation data to predict the global transmission dynamics of human influenza H3N2. PLoS Pathog. 10, e1003932 (2014).

